# Biophysical mechanisms of electroconvulsive therapy-induced volume expansion in the medial temporal lobe: a longitudinal *in vivo* human imaging study

**DOI:** 10.1101/2021.04.19.21255633

**Authors:** Akihiro Takamiya, Filip Bouckaert, Maarten Laroy, Jeroen Blommaert, Ahmed Radwan, Ahmad Khatoun, Zhi-De Deng, Myles Mc Laughlin, Wim Van Paesschen, François-Laurent De Winter, Jan Van den Stock, Stefan Sunaert, Pascal Sienaert, Mathieu Vandenbulcke, Louise Emsell

**Affiliations:** KU Leuven, Leuven Brain Institute, Department of Neurosciences, Neuropsychiatry, Leuven, Belgium; Geriatric Psychiatry, University Psychiatric Center KU Leuven, Belgium; Department of Neuropsychiatry, Keio University School of Medicine, Tokyo, Japan; KU Leuven, Department of Oncology, Gynaecological Oncology, Leuven, Belgium; KU Leuven, Department of Imaging & Pathology, Translational MRI, Leuven, Belgium; KU Leuven, Leuven Brain Institute, Department of Neurosciences, Research Group Experimental Oto-rhino-laryngology, Leuven, Belgium; Noninvasive Neuromodulation Unit, Experimental Therapeutics and Pathophysiology Branch, National Institute of Mental Health, National Institutes of Health, Bethesda, Maryland, USA; Department of Psychiatry and Behavioral Sciences, Duke University School of Medicine, Durham, North Carolina, USA; KU Leuven, Leuven Brain Institute, Department of Neurosciences, Research Group Experimental Neurology, Leuven, Belgium; Department of Radiology, University Hospitals Leuven (UZ Leuven), Leuven, Belgium; Academic Center for ECT and Neuromodulation (AcCENT), University Psychiatric Center, KU Leuven, Kortenberg, Belgium

## Abstract

**Background:** Electroconvulsive therapy (ECT) applies electric currents to the brain to induce seizures for therapeutic purposes. ECT increases gray matter (GM) volume, predominantly in the medial temporal lobe (MTL). The contribution of induced seizures to this volume change remains unclear.

**Methods:** T1-weighted structural MRI was acquired from thirty patients with late-life depression (mean age 72.5±7.9 years, 19 female), before and one week after one course of right unilateral ECT. Whole brain voxel-/deformation-/surface-based morphometry analyses were conducted to identify tissue-specific (GM, white matter: WM), and cerebrospinal fluid (CSF) and cerebral morphometry changes following ECT. Whole-brain voxel-wise electric field (EF) strength was estimated to investigate the association of EF distribution and regional brain volume change. The association between percentage volume change in the right MTL and ECT-related parameters (seizure duration, EF, and number of ECT sessions) was investigated using multiple regression.

**Results:** ECT induced widespread GM volume expansion with corresponding contraction in adjacent CSF compartments, and limited WM change. The regional EF was strongly correlated with the distance from the electrodes, but not with regional volume change. The largest volume expansion was identified in the right MTL, and this was correlated with the total seizure duration.

**Conclusions:** Right unilateral ECT induces widespread, bilateral regional volume expansion and contraction, with the largest change in the right MTL. This dynamic volume change cannot be explained by the effect of electrical stimulation alone and is related to the cumulative effect of ECT-induced seizures.

## Introduction

Electroconvulsive therapy (ECT) is an established effective treatment for severe depression, especially for older adults [1–4]. Although ECT induces multiple generalized seizures by the application of electric currents through the scalp under general anesthesia, ECT is a safe treatment with a low mortality rate [5], and is not associated with long-term cognitive decline in older patients [6, 7] nor with increased risk of developing dementia [8].

Gray matter volume (GMV) increase in the medial temporal lobe (MTL), including the hippocampus and amygdala has been consistently reported following ECT [9–11]. Moreover, ECT has been shown to increase dentate gyrus (DG) volume [12–17], a highly neuroplastic region that may be modulated by applied electric currents and seizures [18]. Notably, DG expansion has been associated with clinical improvement [16, 17, 19], although the clinical relevance of this volume change is unresolved [20–22].

A recent large mega-analysis reported that GMV increase following ECT was not specific to the MTL, but widely distributed across the cortex, with the largest GMV increase in the MTL [23]. Although GMV increase following ECT has been consistently reported, the biophysical mechanism of this phenomenon is still unclear. There are multiple aspects of ECT that could contribute to the observed changes. For example, electrode placement has been associated with the degree of volume change [11, 23, 24]. The number of ECT sessions may also be an important factor, with more ECTs being associated with greater hippocampal volume increase [11]. Notably, another mega-analysis [25] did not find such association. The strength and distribution of the applied electric field (EF) has also been associated with ECT-related GMV increase in the left MTL, but not in the right MTL [26]. In addition to these ECT-related parameters, there is another potential candidate which could have an effect on brain structural change following ECT: seizure. Temporal lobe epilepsy is the most frequent form of focal epilepsy [27]. The hippocampus has a lower seizure threshold compared to other brain regions [28] and the amygdala is considered one of the most susceptible brain regions to seizures [29]. Moreover, reversible peri-ictal MRI signal changes are frequently reported [30–32] particularly in the MTL [33] and are hypothesized to reflect the consequences of ictal hemodynamic and metabolic changes that are associated with vasogenic and cytotoxic edema. However, several studies report an absence of increased visible T2 signal or diffusivity changes following ECT, which argues against edema contributing to volumetric change [34–39]. Whilst epileptic seizures arise against a backdrop of neuropathology which limits the utility of epilepsy as a neurobiological model for ECT, there remain parallels between the two conditions which could provide useful insights into post-ictal structural brain changes following ECT. As far as we know, there is only one diffusion tensor imaging (DTI) study reporting that mean seizure duration was associated with decreased fractional anisotropy, a proxy for white matter (WM) organization, following ECT [40].

From a methodological point of view, most previous studies have focused on ECT-related GMV changes using voxel-based morphometry (VBM) [41], surface-based morphometry (SBM) [15, 42– 44], or region-of-interest (ROI) analysis [11, 12, 35, 36], all of which rely on tissue segmentation. Deformation-based morphometry (DBM) is an alternative way to explore brain structural changes at a macroscopic level [45]. DBM does not rely on tissue segmentation, but utilises information derived from image registration during spatial normalization. Hence, DBM can be used to investigate global volumetric or shape changes regardless of tissue type [45–47]. In addition, DBM is considered more sensitive to WM change than VBM [48], therefore, combining VBM, SBM and DBM could provide new insights into the effect of ECT on brain morphometry. In the present study, we investigate how EF distribution and total seizure duration relate to brain morphometry change following ECT. Based on previous literature [23, 26, 41], we hypothesized that 1) volume expansion would predominantly occur in the MTL GM, and that 2) MTL volume expansion would be associated with EF and cumulative seizure duration.

## Materials and Methods

### Participants

This study was approved by the ethical committee of the Leuven University Hospitals (S5144) and conducted in accordance with the Declaration of Helsinki. Written informed consent was obtained from all participants. Participants were recruited from the inpatient ward at the University Psychiatric Center KU Leuven, Kortenberg, Belgium. Inclusion criteria were a diagnosis of a major depressive disorder according to DSM-IV and age older than 55 years. The diagnoses were confirmed by Mini-International Neuropsychiatric Interview (MINI) assessment. Exclusion criteria were another major psychiatric illness, alcohol or drug dependence, a history of a major neurologic illness (e.g., Parkinson disease, stroke, and dementia) and metal implants precluding MRI. Psychotropic medication was discontinued at least one week before ECT or kept stable from six weeks before ECT and during the ECT course if discontinuation was deemed impossible.

### ECT procedure

ECT was administered twice a week with a constant-current, Thymatron System IV device (Somatics, Venice, FL). Motor and electroencephalographic (EEG) seizures were monitored to ensure adequate duration and quality. Duration of EEG seizure activity was measured by two-channel EEG (Fp1-M1, Fp2-M2, international 10-20 system). All participants in this study were treated with brief pulse (0.5 ms), 900 mA current amplitude, right unilateral (RUL) ECT with dosed at six times the initial seizure thresholds, as determined at the first session. ECT was continued until the patients achieved sustained remission defined by Montgomery-Åsberg Depression Rating Scale (MADRS) score <10 in two consecutive ratings with a one-week interval or until no more improvement was seen. Patients who did not respond to RUL ECT by the sixth session were switched to bitemporal ECT (n = 5). These patients were excluded from this study to ensure sample homogeneity because EF distribution is related to electrode placement [49]. Etomidate (0.2mg/kg) was used for general anesthesia, and succinylcholine (1 mg/kg) was used to induce muscle relaxation.

### MRI data acquisition

Structural MRI data were acquired within one week before and after the last ECT using a 3T Philips Intera scanner with an eight-channel head coil. 3D magnetization prepared rapid gradient echo (MPRAGE) T1-weighted images were acquired: TR = 9.6 ms, TE = 4.6 ms, slice thickness = 1.2 mm, voxel size = 0.98 × 0.98 × 1.2 mm^3^, 182 slices. Diffusion MRI data were acquired prior to ECT in 24 participants using a sagittally acquired echo planar imaging sequence with diffusion-weighting, b=800s/mm^2^, applied along 45 uniformly distributed gradient directions, and included six non-diffusion weighted images, TR/TE = 11000ms/55ms, 68 slices, voxel size 1.98 × 1.98 × 2.2mm^3^.

### Morphometry analysis

The images were visually checked for the presence of any gross motion artifacts or any pathological brain changes preventing further analysis. All pre-processing was conducted using the longitudinal processing pipeline of the Computational Anatomy Toolbox (CAT12) with Statistical Parametric Mapping (SPM)12, including within-subject rigid-body registrations and bias-field corrections. Spatially normalized, modulated tissue-classified images were used to conduct VBM analysis. Each image was smoothed with an 8 mm isotropic full-width at half-maximum (FWHM) Gaussian kernel. In addition, Jacobian maps from each time point were extracted to conduct DBM analysis. Normalized Jacobian maps were smoothed using two different FWHM Gaussian kernels: an 8 mm kernel was used in line with the VBM analysis, and a 2 mm kernel was used to refine the location of ECT-related non-tissue specific volume expansion or contraction. To complement the results from VBM and DBM, we also conducted a surface-based morphometry (SBM) analysis to investigate the effect of ECT on cortical thickness. We used the default pipeline implemented in the CAT12 for SBM. Cortical thickness was estimated using the projection-based thickness method [50]. This algorithm uses tissue segmentation to estimate the WM distance and projects the local maxima to other GM voxels using a neighbor relationship that is described by the WM distance. The vertex-wise thickness measures were resampled and smoothed with a 12 mm geodesic FWHM Gaussian kernel which is the minimum kernel size recommended in the CAT12 manual. Paired t-tests were conducted using SPM12 to detect brain morphometry changes following ECT. The statistical threshold was set at a cluster-level family-wise-error (FWE) corrected p <0.05 with an individual voxel-height threshold of p <0.001. To visually compare the results from the three morphometry analyses, the resulting statistical maps were overlaid on the same surface template.

### Electric field modeling

Each individual finite element head model was constructed using the SimNIBS 3.1.2 toolbox (https://simnibs.github.io/simnibs/build/html/index.html). We used a “headreco” option, which used CAT12/SPM12 for the segmentation of each T1-weighted image and for the creation of the tetrahedral finite element mesh. The head model consisted of five tissue compartments: skin, skull, CSF, GM, and WM, with assigned isotropic conductivities of 0.465 S/m, 0.010 S/m, 1.654 S/m, 0.276 S/m, and 0.126 S/m, respectively [51]. Rectangular electrodes of 4 × 5 cm were placed over the C2 and FT8 EEG sites according to the international 10-20 system to simulate the RUL electrode placement [26]. EF simulation was conducted with an electric current of 1 mA. After normalization to MNI space, the resultant images were multiplied by 900 to be scaled to the current amplitude of the Thymatron device. These processes resulted in subject-specific voxel-wise EF magnitude maps in MNI space. In the model, we assumed that the applied current is static, with the maximum amplitude of the pulse the same as [26].

We also created individual finite element head model incorporating diffusion MRI data to improve the estimation of the EF distribution in 24 participants. The diffusion tensors were estimated using a non-linear least squares approach based on [52] following motion and distortion correction in ExploreDTI [53]. We used dwi2cond command implemented in the SimNIBS, which depends on FMRIB Software Library (FSL) and uses a non-linear registration based on FSL fnirt to transform the tensor imaging to the space of the T1-weighted images. WM anisotropy was incorporated based on conductivity tensors derived from diffusion tensor modelling using a volume normalized mapping approach [54] with maximum eigenvalue of 2 and maximum ratio between eigenvalue of 10. One dataset was excluded because of registration failure.

### Contributing factors to morphometry change following ECT

To illustrate the voxel-wise distribution of the estimated EF and volume change following ECT, mean images of EF distribution and volume change from all participants were created.

A multiple regression analysis was conducted to identify which ECT-related parameters were associated with the observed brain volume change following ECT. In this regression model, the percentage volume change in the peak cluster based on T-value in the DBM analysis with 2 mm smoothing kernel was included as a dependent variable. We used the DBM result because ECT could affect both GM and WM, which can be captured by DBM simultaneously. To investigate the effect of seizure properties and electrical stimulation on ECT-related brain volume change, total EEG seizure duration measured by two-channel EEG during the ECT procedure, and the estimated EF in the peak cluster in the DBM analysis were included as independent variables in the regression model. Duration of EEG seizure at the first session was excluded when calculating the total EEG seizure duration because the first session is used to measure each individual’s seizure threshold. Missing values of the seizure duration were imputed using the mean value in each participant. The number of ECTs, which has been associated with hippocampal volume increase following ECT [11], was also included in the model. However, the total seizure duration and number of ECTs are highly correlated (r = 0.68, p <0.001). Thus, we included residuals of the number of ECTs after regressing out the effect of the total seizure duration to account for the effect of the number of ECTs, which is not explained by the seizure duration. Finally, we conducted a correlation analysis investigating associations between EF and the effect sizes of regional volume expansion in each ROI defined by the Hammers brain atlas [55] (supplementary table 1).

**Table1.** Clinical characteristics of the participants

|  | ECT patients |
| --- | --- |
| Number | 30 |
| Age (years) | 72.5 (7.9) |
| Female | 19 (63%) |
| Psychotic features | 15 (50%) |
| Late-onset depression | 15 (50%) |
| ECT parameters |  |
| Number of ECTs | 11 (23) |
| Total EEG seizure (seconds) | 490 (140) |
| MADRS at TP1 | 36 (8) |
| MADRS at TP2 | 8 (10) |
| Clinical response, n (%) | 27 (90%)* |
| Clinical remission, n (%) | 25 (83%)* |

Continuous variables are expressed as mean (standard deviation).
\*Note that we excluded patients who did not respond to RUL ECT and switched to bilateral ECT.

## Results

In the current study, we included 30 older adults with late life depression (age: 72.5 ± 7.9, female: 19 (63%)) who received an average of 11 ± 3 right unilateral ECT sessions (range: 4–16). Clinical characteristics of the participants and ECT parameters are presented in Table 1.

### Morphometry analysis

VBM showed cortical and subcortical GMV increase, bidirectional WM volume (WMV) change, and CSF decrease following ECT. The WMV decrease occurred mainly adjacent to the regions of GMV increase, and the WMV increase occurred mainly in the periventricular regions (Figure 1). When using an 8-mm smoothing kernel, DBM showed widespread volume expansion in both GM and WM with corresponding contraction in adjacent CSF compartments. DBM with 2-mm smoothing kernel revealed that the volume expansion was mainly located in the GM, and that limited WM volume change was observed (Figure 2). The largest volume expansion was identified in the right MTL surviving voxel-level FWE-correction p <0.05 (supplementary figure 1). SBM analysis showed widespread cortical thickness increase bilaterally (Figure 3A). To visually compare the results of the morphometry analyses, all morphometry results were overlaid on the same surface template (Figure 3).

**Figure 1.**
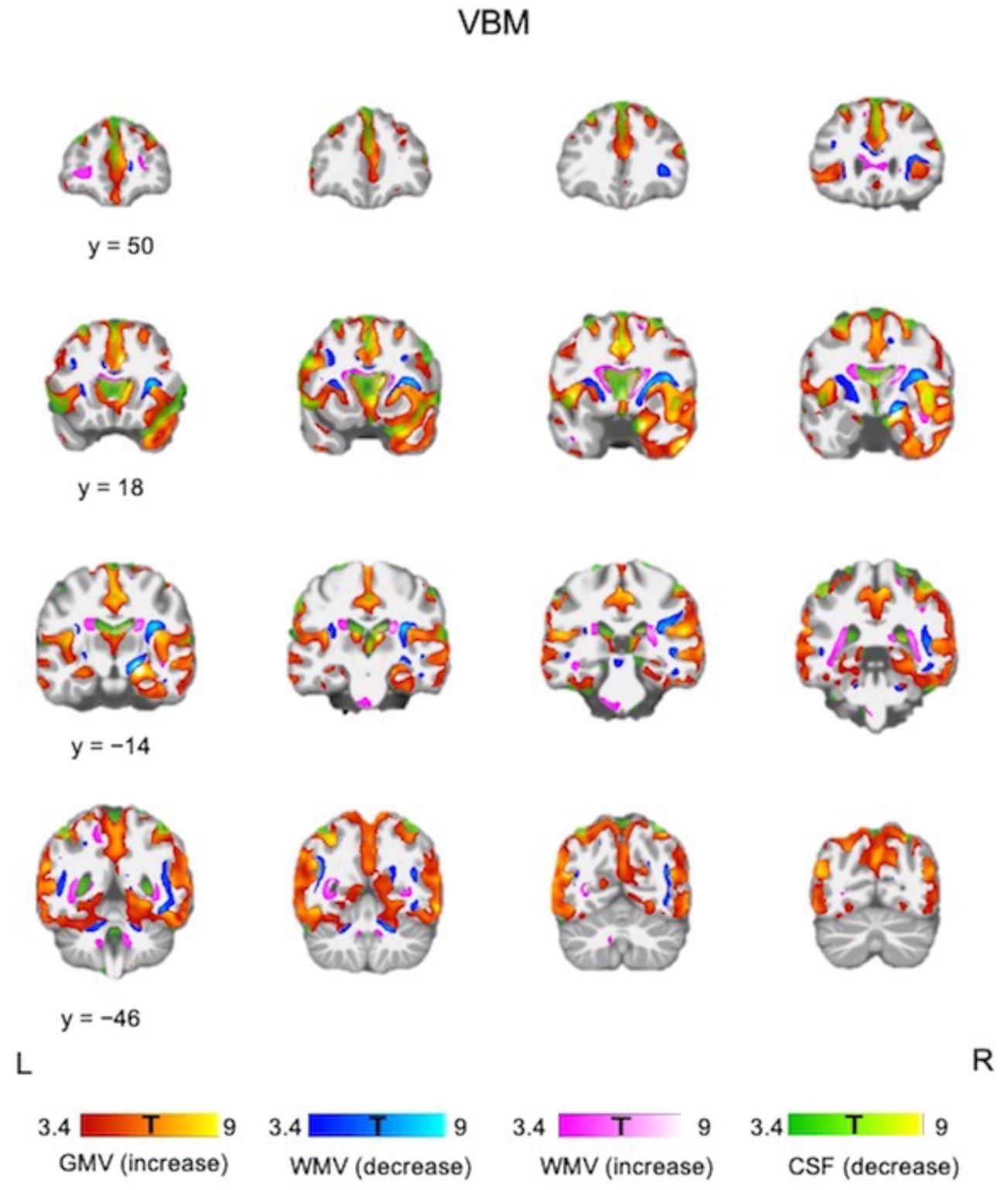
Tissue-specific volume change following ECT. VBM showed bilateral GMV increase and CSF decrease. WMV increase was observed in the periventricular regions, and WMV decrease was observed adjacent to the regions of GMV increase. Each color bar at the bottom of the figure represents the T-values derived from the VBM results.

**Figure 2.**
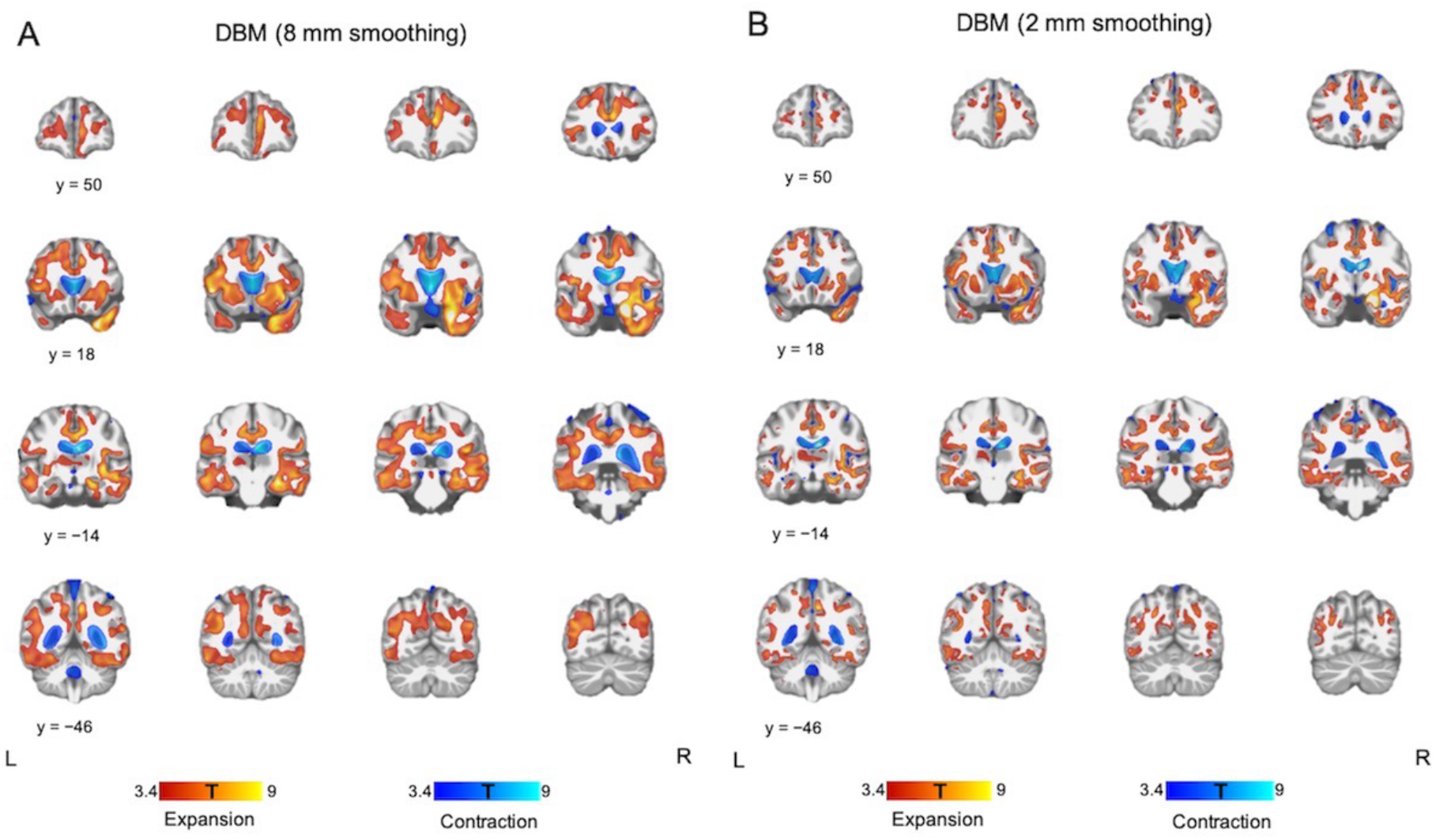
Global volumetric change following ECT. (A) DBM using images smoothed with an 8 mm kernel showed bilateral volume expansion in GM and WM with contraction in the CSF compartments. (B) DBM using images smoothed with a 2 mm kernel revealed that volume expansion was mainly located in the GM. Each color bar at the bottom of the figure represents the T-values derived from the DBM results.

**Figure 3.**
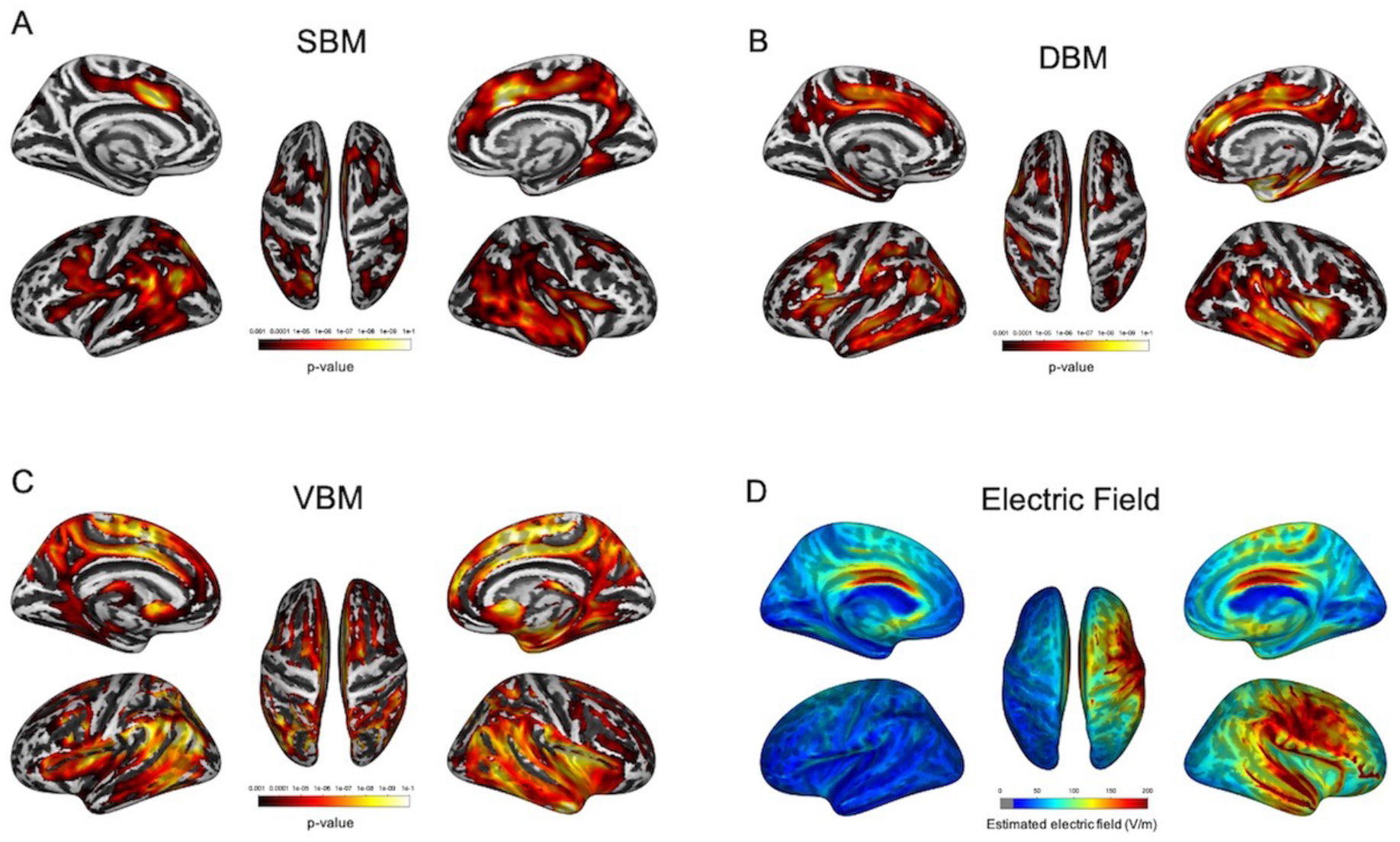
Results of morphometry analyses following ECT and estimated electric field strength. (A–C) SBM and DBM identified almost identical brain regions, whereas VBM showed more widespread volumetric change pattern following ECT. Each color bar represents p-value in the whole-brain analysis. (D) Figure 3D shows the mean image of the estimated electric field strength in all participants. The color bar represents electric field strength (V/m).

### Voxel-wise electric field estimation and brain shape change following ECT

Mean voxel-wise EF distribution and volume expansion following ECT are presented in Figure 4, and the EF distribution is also overlaid on the surface template (Figure 3D). Top-ranked brain regions exposed to the highest EF and brain regions showing the largest volume expansion are presented in supplementary table 2. There was a correlation between the EF and effect size of regional volume expansion (Spearman’s rho = 0.27, p = 0.04), but no correlations when analyzing each hemisphere separately (supplementary figure 2). The EF strongly correlated with the distance from the brain regions near the electrodes (i.e., right precentral gyrus and right superior temporal gyrus) (supplementary figure 3).

**Table 2.** ECT parameters associated with percentage volume expansion in the right MTL.

| | B | SE | $\beta$ | p-value |
| --- | --- | --- | --- | --- |
| Total EEG seizure duration | 0.01 | 0.006 | 0.39 | 0.039 |
| Electric field | 0.11 | 0.09 | 0.24 | 0.20 |
| Number of ECTs (residuals)* | 0.38 | 0.38 | 0.18 | 0.33 |

B: non-standardized coefficient; SE: standard error; $\beta$ : standardized coefficient
\* The number of ECTs (residuals) is the information about the number of ECTs that is not explained by the total EEG seizure duration, hence due to the high correlation ( $r = 0.68$ , $p < .001$ ) all information about the number of ECTs is included in this regression model.

**Figure 4.**
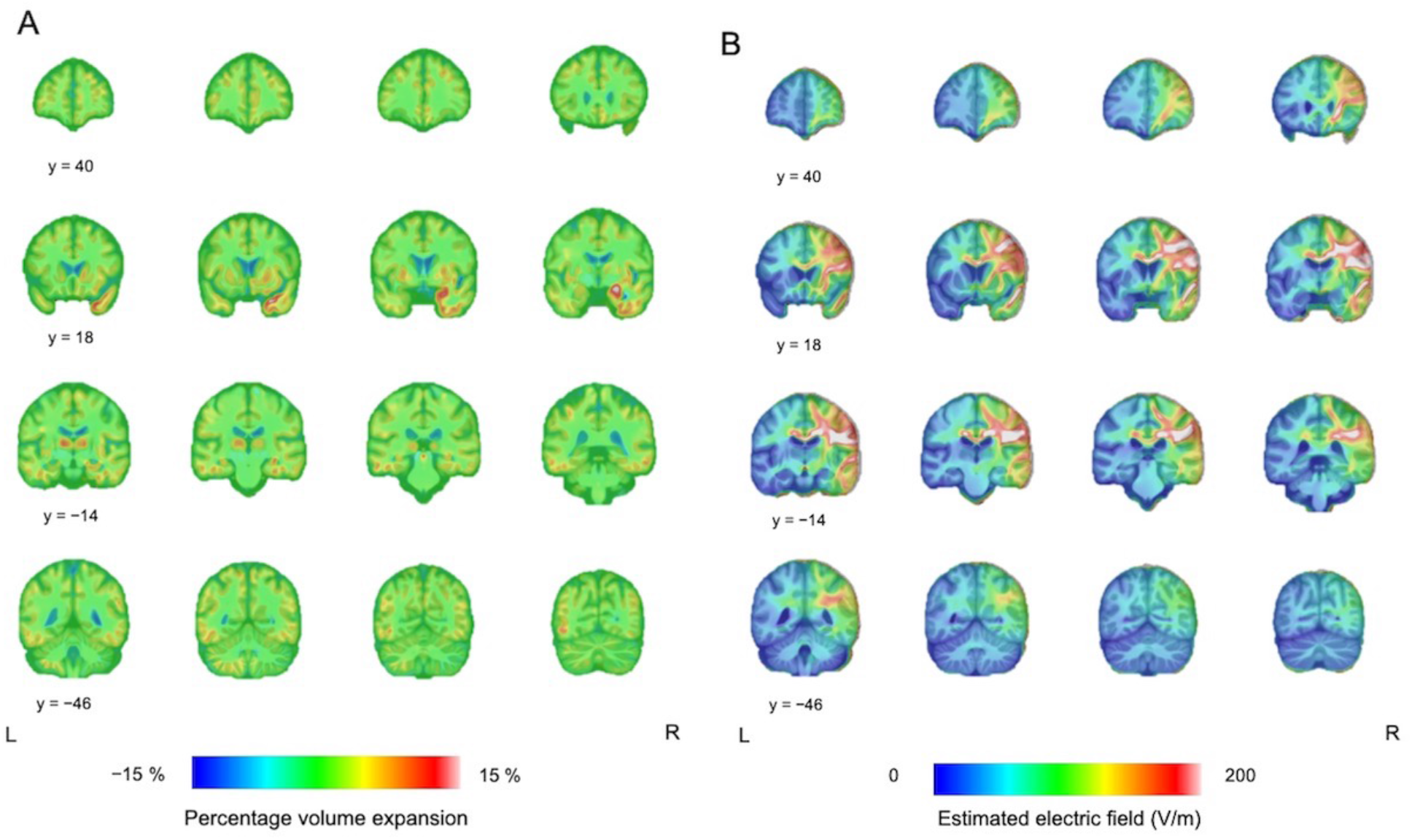
Different patterns of the volume change following ECT and estimated electric field strength. (A) Figure 4A shows the mean image of the DBM results. The largest volume expansion was observed in the right medial temporal lobe. The color bar represents percentage volume expansion. (B) Figure 4B shows the mean image of the estimated electric field. The color bar represents electric field strength (V/m).

### Association between ECT-related parameters and volume expansion

In the multiple regression model of the full sample, the total EEG seizure duration was associated with the percentage volume expansion in the right MTL (p = 0.039, Table 2). In the subgroup (n = 23) incorporating DTI derived anisotropy data to improve the EF model, the result of the regression analyses was similar: the total EEG seizure duration was associated with the percentage volume expansion in the right MTL (p = 0.045, supplementary table 3).

## Discussion

Our results provide evidence of dynamic volume changes following ECT, suggesting that GM volume expansion asserted a mass effect on adjacent tissue with compensatory volume decrease in CSF-filled spaces. The effect on WM was less pronounced, and the volumetric changes observed with VBM could be interpreted as a displacement based on the DBM results. Moreover, the largest volume expansion in the right MTL did not correlate with the EF distribution, but with the total seizure duration over the ECT course.

### Differential effect of ECT on gray and white matter

In this study, DBM revealed that brain deformation changes following ECT were observed mainly in the GM and CSF compartments, but less change was identified in the WM. This discrepancy is interesting as it argues against a uniform effect of ECT on brain tissue. Although the effect of ECT on WM has been reported in several DTI studies [40, 56–58], these results were inconsistent, and some were contradictory. Some may argue that T1-weighted imaging lacks sensitivity for detecting WM changes. However, previous DBM analyses have reported epilepsy-related [59] and chemotherapy-related [60] WM changes. Therefore, the effect of ECT on WM might be too small to detect using DBM analysis or ECT might not cause macroscopic WM change (e.g., axonal swelling). Although our VBM analysis revealed a bidirectional WMV change pattern, these changes were observed mainly next to expanded GM or contracted CSF-filled spaces (Figure 1). In addition, there were significant correlations among the degree of GMV increase, WMV increase/decrease, and CSF decrease (supplementary figure 4). Taken together, these findings suggest that the WMV changes detected by VBM may reflect WM displacement due to adjacent expanded GM.

In line with the literature [11, 12, 15, 23, 35, 41–44, 61], all morphometry analyses consistently found both GM volume and cortical thickness increase following ECT, with the largest change in the right MTL. Multiple mechanisms may give rise to this GM expansion, including neuroplastic changes, changes in intracellular and extracellular fluid compartments (e.g., due to edema), immunological activation and altered cerebral blood flow [18, 62]. In general, more than 80% of the cortical volume is made up of neuropil (i.e., the axons, dendrites, synapses, glial cell processes and microvasculature), and the rest is made up of neuronal cell bodies, blood vessels and glial cell bodies [63]. The number of neurons in the cortex is considered to remain largely constant [64], therefore, cortical volumetric expansion and cortical thickness increase may reflect an increase in dendrites and synapses [63]. Neurogenesis has been postulated as a potential mechanism underlying volume increase in the hippocampus [18], but the magnitude of the MTL volume expansion (>5%) and cortical extent of GM change suggests other, additional neuroplastic mechanisms also contribute to the volume increase. Synaptic plasticity might be accompanied by growth of both capillaries and glia [66], perhaps to support the increased energy demand of new synapses [67]. Indeed, electroconvulsive seizures (ECS), an animal model of ECT, has been shown to alter the number, morphology, and activation of glial cells [68], including astrocytes [69]. Interestingly, astrocytic activation may change the MRI T1-relaxation time manifesting as GMV increase [70], and vascular changes could also affect GMV estimation because of the relatively similar relaxation time constants between arterial blood and GM [71].

Another consequence of vascular and immunological changes is edema. Volume increase in the periictal phase following epileptic seizure has been detected by other MRI sequences (e.g., T2-weighted images) and attributed to cellular edema [72]. However, DTI studies have consistently reported decreased water diffusivity in the MTL following ECT [36–38]. Whilst reduced mean diffusivity could theoretically reflect acute cytotoxic edema, a recent multi-contrast quantitative MRI study reported that GMV increase following ECT was not paralleled by an increase in proton density, a proxy for tissue water content. These findings suggest that neither vasogenic nor cytotoxic edema are likely to be the primary contributor to MTL volume increase following ECT [39].

### Effect of electrical stimulation on regional volume change following ECT

We found that regional EF strength, which is a proxy for the effect of direct electrical stimulation rather than the seizure activity, was the highest in the brain regions near the stimulus electrodes (i.e., GM of the right pre/post central gyrus and the right superior temporal gyrus with the greatest magnitude in the adjacent WM), in line with a previous study [49]. The regional EF was strongly correlated with the Euclidean distance from these brain regions (supplementary figure 3), but not with the regional volume change when analyzing the ROIs in each hemisphere separately. The right pre/post central gyrus did not show statistically significant volume or thickness change even with the exposure to the highest EF. Indeed, the spatial distribution of the volume changes are more bilaterally symmetric compared to the lateralized EF distribution induced by RUL ECT (Figure 3), although the volume expansion near the hippocampus appear more lateralized to the right hemisphere (Figure 4). Interestingly, MTL volumetric increase has been reported following transcranial magnetic stimulation in several studies [72, 73], but not all [74], and it has also been reported following vagus nerve stimulation [75]. These results suggest that the topography of the applied electrical field alone cannot explain the regional volume change following ECT, especially the largest volume expansion in the right MTL.

Our results and arguments do not fully contradict a recent mega-analysis (which also included data from the present study) [26]. We replicated the significant correlation between the EF and left hippocampal volume expansion (p = 0.04), and a correlation between the EF and left amygdala volume expansion (p = 0.056) (data not shown) despite several methodological differences between our analyses.

### Effect of seizure on regional volume change following ECT

In the present study, the right MTL volume expansion was associated with the total EEG seizure duration, i.e., the sum of the seizure duration per session. ECT-induced seizures are focal to bilateral tonic-clonic seizures, involving certain specific cortical and subcortical brain regions, while other brain regions are less affected [76]. Moreover, RUL ECT-induced seizures are not always symmetrical, but can be asymmetrical with higher EEG amplitude on the right-side during seizures [77]. In this context, the origin and propagation pattern of ECT-induced seizure is relevant. The MTL or precentral gyrus may be the origin of ECT-induced seizure because of their lower seizure threshold relative to other brain regions [28]. We found large volume expansion in the right amygdala, which is in line with a previous mega-analysis [23]. Interestingly, the amygdala is often chosen as a target for kindling in preclinical research due to its susceptibility for seizure generation [78]. One animal study including 25 cats reported that bilateral ECT-induced seizures initiated in the sensorimotor cortex resulted in the largest discharge in the hippocampus. Moreover, the hippocampal seizure activity continued longer than in other brain regions and propagated to functionally connected regions, such as the amygdala [79]. Another study which investigated epileptic seizure propagation from the hippocampus reported intense activation of the hippocampus resulting from re-entrant seizure activity, and hippocampal activity throughout the course of the seizure in contrast to the cortical structures [80]. Our results suggest that the estimated EF was large enough to elicit neuronal depolarization in almost all brain regions (supplementary material), therefore other brain regions or several brain regions could be the origin(s) of ECT-induced seizure.

The relationship between seizures and brain volumetric change is complicated. There are several differences between ECT-induced seizures and disease-related epileptic seizures. For example, the effect of epileptic seizures on brain structure may vary across brain regions and includes volume reduction in the MTL and thalamus [81], and volume increase in the amygdala [82], which may normalize following successful antiepileptic treatment [83]. According to preclinical studies, epileptic seizures result in the generation of new neurons characterized by structural and functional abnormalities [84], which is in contrast with ECS-treated animals, where no new-born neurons in the hilus (ectopic) were observed [85, 86]. ECS induces sprouting of the granule cell mossy fiber pathway in the hippocampus without generation of abnormal connectivity and does not lead to the appearance of spontaneous epileptic activity. ECS-induced sprouting occurs in the absence of neuronal loss, indicating that sprouting is not a compensatory response to cell death [87].

### Limitations

Measuring the applied current and seizure duration, and investigating their relationships is not trivial. Firstly, we measured seizure duration using two-channel EEG in a clinical setting. Secondly, we used the summed seizure duration to assess the cumulative effect of seizures across the whole ECT course. This is because the seizure duration decreased later in the course of ECT, which may be due to increased seizure threshold (supplementary material). This heterogeneity was not accounted for in our models. In addition, whilst we attempted to statistically disentangle the effects of total seizure duration from the number of ECT sessions because they are highly correlated, this relationship remains complicated as the number of ECTs also correlates with opposing seizure-related parameters (supplementary figure 5). As in the mega-analysis [26], the EF model accounted for spatial aspect (including current amplitude) but not temporal information of the pulse train (e.g., pulse width and polarity, frequency, and number of pulses). Parameters of the temporal pulse sequence such as stimulus train frequency and duration can significantly affect seizure duration and postictal suppression [88]. These parameters are typically reported as part of an aggregate measure of dosing, i.e., stimulus charge. Future studies should consider the effects of individual stimulus parameters on seizure expression and brain volume change. There is also a need to investigate the role of the EF on the seizure initiation and subsequent propagation.

## Conclusion

ECT induced widespread GM expansion with volume contraction in the CSF compartments. The largest volume expansion in the right MTL could not be explained by the effect of electrical stimulation alone and was associated with total seizure duration during a course of therapy.

## Data Availability

The data that support the findings of this study are available on reasonable request from the corresponding author. The data are not publicly available because this could compromise the privacy of the research participants.

## Acknowledgements

This study was supported by the Research Foundation Flanders (FWO) project G074609 (M. Vandenbulcke), G0C0319N (M. Vandenbulcke, F. Bouckaert, L. Emsell), Research Grant Old Age Psychiatry UZ Leuven R94859 (M. Vandenbulcke), KU Leuven C24/18/095 (M. Vandenbulcke, F. Bouckaert, J. Van den Stock, L. Emsell) and KU Leuven Sequoia Fund.

A. Takamiya has received grants from Keio University Medical Science Fund and Kanae Foundation for the Promotion of Medical Science to study in Belgium. J.Blommaert is an aspirant researcher for the Research Foundation Flanders (FWO, grant no. 11B9919N). Z.-D. Deng is supported by the Intramural Research Program of the National Institute of Mental Health, National Institutes of Health (ZIAMH002955).

## Author contributions

**Akihiro Takamiya:** Conceptualization, Methodology, Formal analysis, Investigation, Writing - Original Draft, Visualization

**Filip Bouckaert:** Conceptualization, Investigation, Writing - Review & Editing, Supervision, Funding acquisition

**Maarten Laroy:** Methodology, Writing - Review & Editing

**Jeroen Blommaert:** Methodology, Writing - Review & Editing

**Ahmed Radwan:** Methodology, Writing - Review & Editing

**Ahmad Khatoun:** Methodology, Writing - Review & Editing

**Zhi-De Deng:** Methodology, Writing - Review & Editing

**Myles Mc Laughlin:** Methodology, Writing - Review & Editing

**Wim Van Paesschen:** Writing - Review & Editing

**François-Laurent De Winter:** Investigation, Writing - Review & Editing

**Jan Van den Stock:** Writing - Review & Editing

**Stefan Sunaert:** Writing - Review & Editing

**Pascal Sienaert:** Writing - Review & Editing

**Mathieu Vandenbulcke:** Conceptualization, Writing - Review & Editing, Supervision, Project administration, Funding acquisition

**Louise Emsell:** Conceptualization, Methodology, Investigation, Writing - Review & Editing, Supervision, Project administration, Funding acquisition

