## supplementary material for "Biophysical mechanisms of electroconvulsive therapy-induced volume expansion in the medial temporal lobe: a longitudinal *in vivo* human imaging study"

Supplementary Table 1. Brain regions determined by the Hammers Atlas.

| |
|---|
| Hippocampus |
| Amygdala |
| Anterior Medial Temporal Lobe |
| Anterior Lateral Temporal Lobe |
| Ambient and Parahippocampal Gyri |
| Superior Temporal Gyrus |
| Inferior Middle Temporal Gyri |
| Fusiform Gyrus |
| Insula |
| Lateral Occipital Lobe |
| Anterior Cingulate Gyrus |
| Posterior Cingulate Gyrus |
| Middle Frontal Gyrus |
| Posterior Temporal Lobe |
| Inferior Lateral Parietal Lobe |
| Caudate Nucleus |
| Accumbens Nucleus |
| Putamen |
| Thalamus |
| Pallidum |
| Precentral Gyrus |
| Gyrus Rectus |
| Orbito-Frontal Gyri |
| Inferior Frontal Gyrus |
| Superior Frontal Gyrus |
| Postcentral Gyrus |
| Superior Parietal Gyrus |
| Lingual Gyrus |
| Cuneus |

Supplementary Table 2. Top-ranked brain regions exposed to the highest EF and brain regions showing the largest volume expansion.

| Electric field (EF) | | | | Volume expansion | | | |
|---|---|---|---|---|---|---|---|
| rank | side | Brain regions | EF (V/m) | rank | side | Brain regions | Effect size (d) |
| 1 | right | Precentral gyrus | 148.2 | 1 | right | Hippocampus | 1.92 |
| 2 | right | Postcentral gyrus | 143.6 | 2 | right | Amygdala | 1.83 |
| 3 | right | Superior temporal gyrus | 127.4 | 3 | right | Anterior medial temporal lobe | 1.77 |
| 4 | right | Inferior lateral parietal lobe | 127.3 | 4 | right | Inferior middle temporal gyrus | 1.71 |
| 5 | right | Middle frontal gyrus | 127.1 | 5 | right | Insula | 1.65 |
| 6 | right | Inferior middle temporal gyrus | 126.5 | 6 | right | Anterior cingulate gyrus | 1.51 |
| 7 | right | Anterior lateral temporal lobe | 122.2 | 7 | right | Posterior cingulate gyrus | 1.36 |
| 8 | right | Inferior frontal gyrus | 118.5 | 8 | right | Ambient and parahippocampal gyrus | 1.34 |
| 9 | right | Nucleus accumbens | 118 | 9 | right | Putamen | 1.27 |
| 10 | right | Pallidum | 107 | 10 | left | Inferior frontal gyrus | 1.25 |

Supplementary Table 3. ECT parameters associated with percentage volume expansion in the right MTL in participants with additional diffusion tensor imaging derived anisotropy modelling

| | B | SE | β | p-value |
|---|---|---|---|---|
| Total EEG seizure duration | 0.01 | 0.006 | 0.45 | 0.045 |
| Electric field | 0.12 | 0.08 | 0.32 | 0.15 |
| Number of ECTs (residuals) | 0.35 | 0.40 | 0.18 | 0.40 |

B: non-standardized coefficient; SE: standard error; β: standardized coefficient

**Supplementary Figure 1. Results of the whole-brain DBM analysis with a strict significance threshold.** Given the number of participants in this study (i.e., 30 participants), a whole-brain significance threshold of a voxel-level FWE-corrected p <0.05 may be too conservative. However, the right MTL volume expansion was still identified at this threshold.

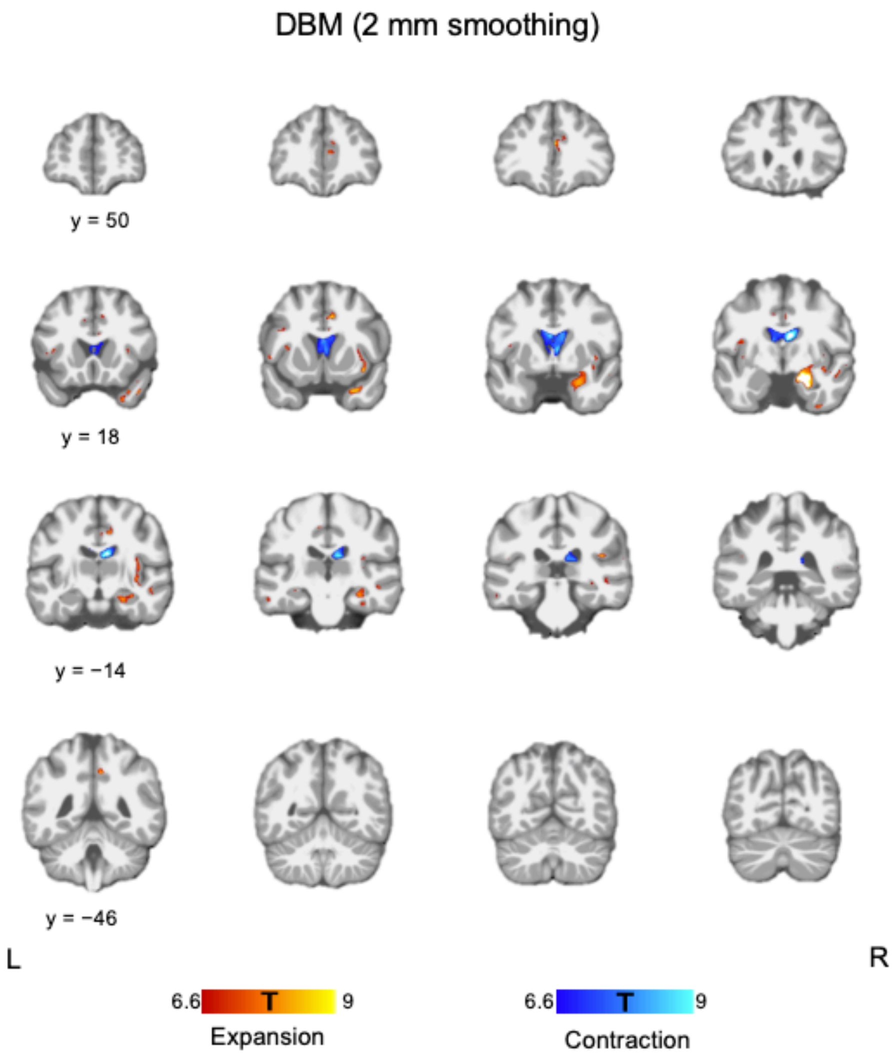

**Supplementary Figure 2**. Electric field distribution and regional volume change following ECT. (a) There was a weak correlation between EF and the effect size of regional volume expansion when including all regions-of-interest in both hemispheres (Spearman's rho = 0.27, p = 0.04). (b) There were no correlations between them when analyzing each hemisphere separately (right: Spearman's rho = −0.03, p = 0.88; left: Spearman's rho = 0.03, p = 0.88). Each region-of-interest (ROI) was defined by the Hammers brain atlas (Supplementary table 1) i.e. each point on the scatter plot represents a different ROI. Y-axis represents the effect size of volume changes (Cohen's d).

(a)

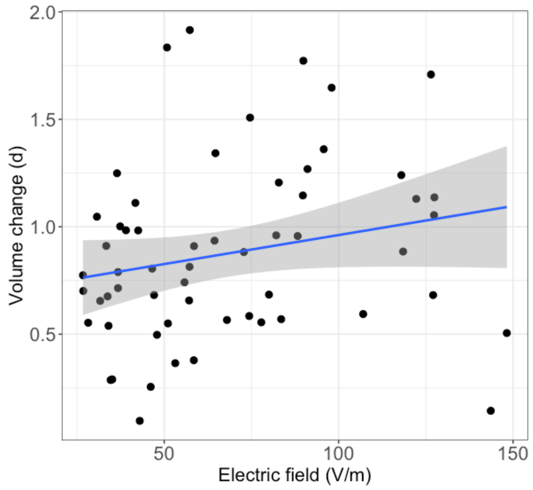


(b)

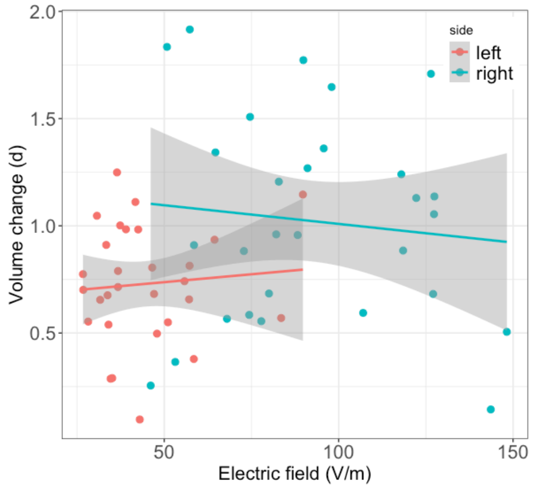

**Supplementary Figure 3**. Electric field distribution and Euclidean distance from brain regions near the stimulus electrodes. (a) There was a strong correlation between EF and Euclidean distance from the right precentral gyrus (PreCG) (r = −0.76, p <0.001). The correlations were significant when analyzing each hemisphere separately (right: r = −0.58, p <0.001; left: r = −0.78, p <0.001). (b) There was a strong correlation between EF and Euclidean distance from the right superior temporal gyrus (STG) (r = −0.74, p <0.001). The correlations were significant when analyzing each hemisphere separately (right: r = −0.37, p = 0.05; left: r = −0.47, p = 0.009). Each region-of-interest was defined by the Hammers brain atlas (Supplementary table 1).

(a)

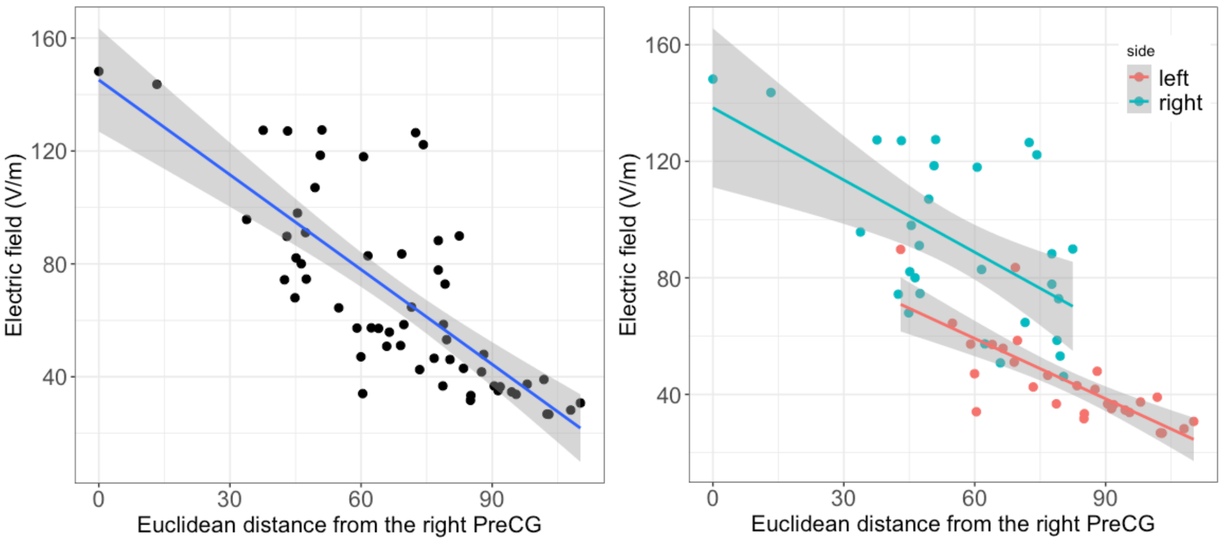


(b)

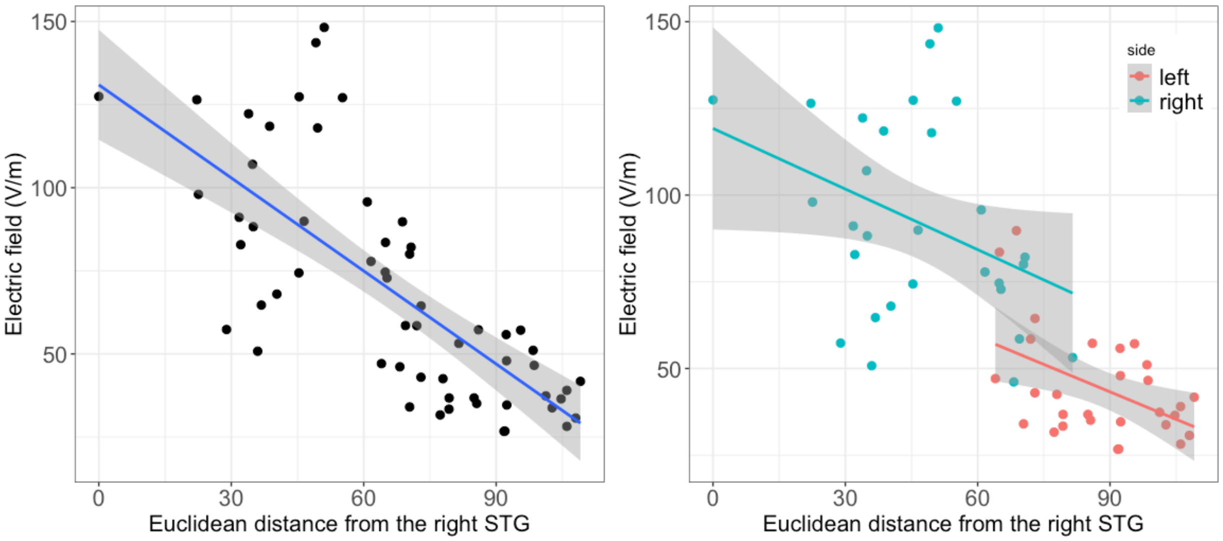

**Supplementary Figure 4. Relationships among tissue-specific volume changes. (a)** GMV increase and WMV decrease (Pearson's r = −0.71, p <0.001), (b) WMV increase and WMV decrease (Pearson's r = −0.46, p = 0.010), and (c) WMV increase and CSF decrease (Pearson's r = −0.60, p <0.001) in the peak regions were significantly correlated with each other. The MNI coordinates in each peak region is the following: GMV increase: (20, −9, −20); WMV decrease: (20, −12, −10); WMV increase: (15, 0, 15); and CSF decrease: (10, −10, 20).

(a)

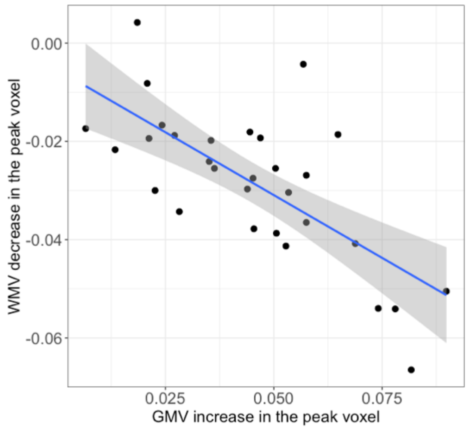


**(b)**

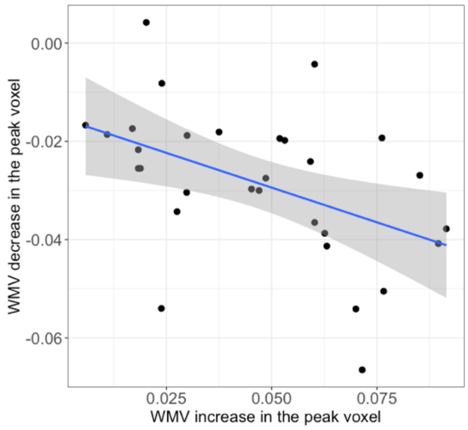


**(c)**

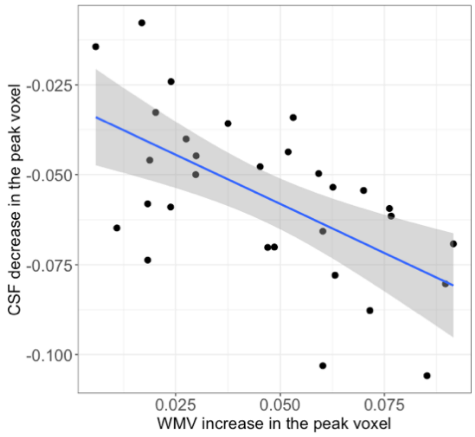

**Supplementary Figure 5**. Relationships between the number of ECTs, seizure duration, and stimulus charge. (a) The number of ECTs was positively correlated with total seizure duration (Pearson's r = 0.68, p <0.001). (b) The number of ECTs was negatively correlated with mean seizure duration (Pearson's r = −0.51, p = 0.004). (c) The number of ECTs was positively correlated with mean stimulus charge (Spearman's rho = 0.44, p = 0.01). (d) Mean stimulus charge was negatively correlated with mean seizure duration (Spearman's rho = −0.48, p = 0.007).

These results could be interpreted in the context of the anticonvulsant effect of ECT (Sackeim, 1999). Although we did not measure seizure threshold during the ECT course, seizure threshold might increase with the number of ECT sessions and therefore clinicians may need to increase the stimulus dose to induce seizures. Higher stimulus charge relative to each individual's seizure threshold produced shorter seizures (Frey et al., 2001).

(a)

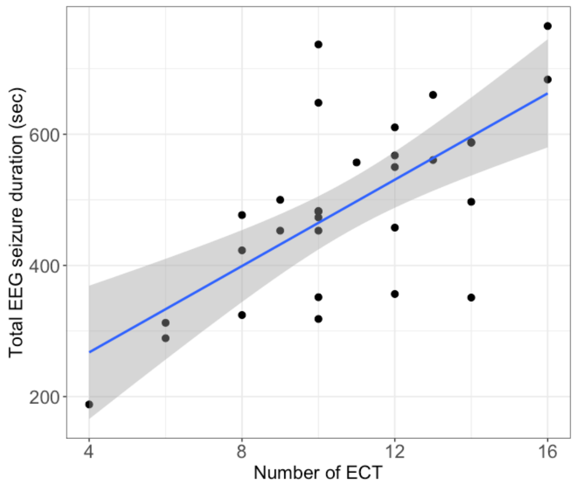


(b)

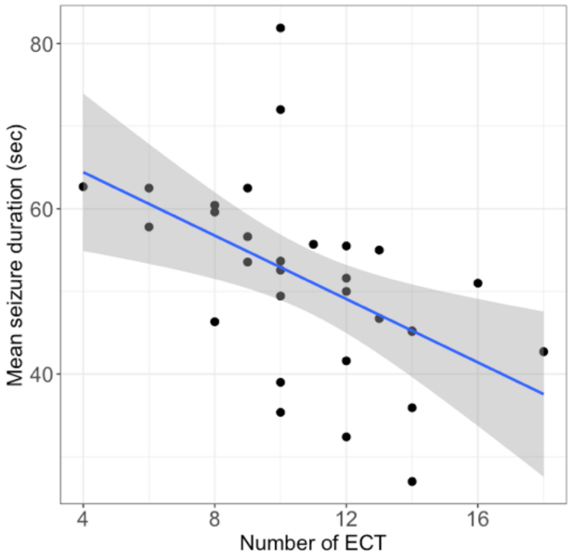


(c)

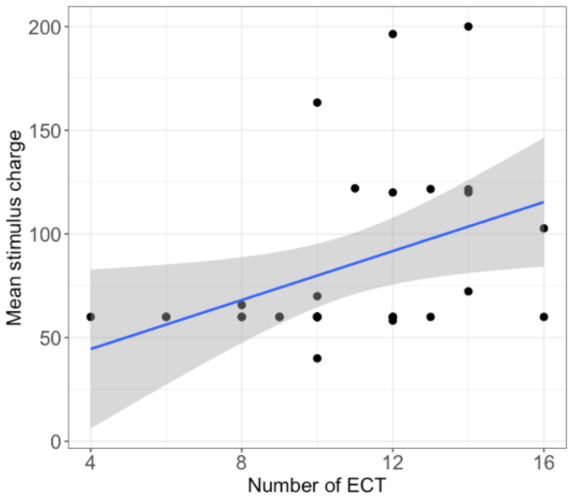


(d)

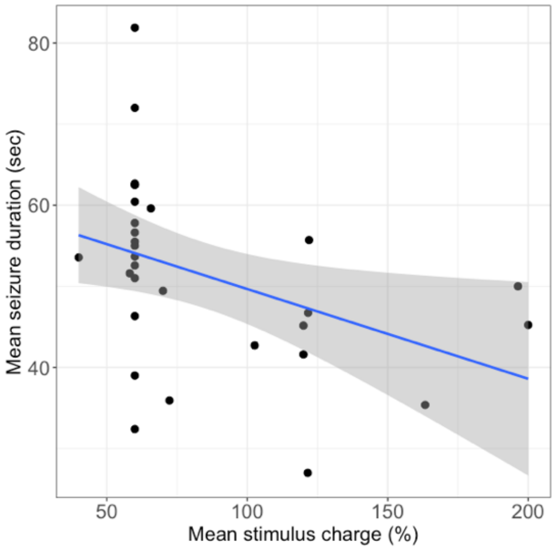
